# Delta-9-tetrahydrocannabinol and cannabidiol in psychosis: A balancing act of the principal phyto-cannabinoids on human brain and behavior?

**DOI:** 10.1101/2021.05.17.21257345

**Authors:** Suhas Ganesh, Jose Cortes-Briones, Ashley M. Schnakenberg Martin, Patrick D Skosnik, Deepak C D’Souza, Mohini Ranganathan

**Author notes:** Corresponding Author Corresponding Author Contact Information: Mohini Ranganathan, Associate Professor, Department of Psychiatry, Yale University School of Medicine, VA Connecticut Healthcare System/116A, 950 Campbell Ave, West Haven, CT 06516.

## Abstract

Delta-9-tetrahydrocannabinol (THC) and cannabidiol (CBD) are the principal phyto-cannabinoids in the cannabis plant. The differential and possibly antagonistic effects of these compounds on specific brain and behavioral responses, and the mechanisms underlying their effects have generated extensive interest in pre-clinical and clinical neuroscience investigations. In this double-blind randomized placebo-controlled counterbalanced human laboratory experiment, we examined the effects of three different dose ratios of CBD: THC (1:1, 2:1 and 3:1) on ‘neural noise’, an electrophysiological biomarker of psychosis known to be sensitive to cannabinoids as well as subjective and psychotomimetic effects. Interestingly, the lowest CBD:THC ratio (1:1) resulted in maximal attenuation of both THC induced psychotomimetic effects (PANSS positive - ATS = 7.83, df = 1, p_corr_ = 0.015) and neural noise (ATS = 8.83, df = 1, p_corr_ = 0.009) with an inverse-linear dose response relationship. Further, in line with previous studies, addition of CBD did not reduce the subjective experience of THC induced “high” (p > 0.05 for all CBD doses).

These novel results demonstrate that CBD attenuates THC induced subjective and objective effects relevant to psychosis- but in a dose/ratio dependent manner. Given the increasing global trend of cannabis liberalization and application for medical indications, these results assume considerable significance given the potential dose related interactions of these key phyto-cannabinoids.

## Introduction

For millennia, *Cannabis sativa*, the cannabis plant, has been used for medicinal and recreational purposes (1) and is one of the most widely used substances today (2, 3). Cannabis contains over 500 distinct chemical compounds and ∼120 phyto-cannabinoids (4) including Δ9-tetrahydrocannabinol (THC), responsible for the psychoactive effects of cannabis, and cannabidiol (CBD), investigated for its therapeutic effects (5, 6).

The acute effects of cannabis include a wide array of subjective, behavioral, and physiological effects including euphoria, relaxation, increased appetite, anxiolysis or increased anxiety, tachycardia, altered spatial and time perception, paranoia, conceptual disorganization, and hallucinations (7-9).

A large body of evidence supports an association between cannabis exposure and psychosis outcomes ranging from acute mild psychosis-like symptoms to the development of a frank psychotic disorder. While data supporting the association between cannabis and psychotic disorders come from epidemiological studies and clinical observations, human laboratory studies (HLS) have complemented this literature to provide temporally related evidence of an association between cannabinoids and psychosis symptoms. Importantly, HLS also permit the examination of underlying mechanisms and interactions between cannabinoids that are not feasible in epidemiological studies. Over the past several decades, HLS have carefully characterized the acute dose-related subjective, cognitive, physiological, neuroendocrine, and psychophysiological effects of THC (10-16). These experimental studies demonstrate that THC administration reliably induces transient psychotic-like symptoms (10, 11), cognitive deficits (11-13), and psychomotor impairments (14, 15).

Unlike THC, CBD has no psychotomimetic or rewarding effects but appears to have sedative and anxiolytic properties (17-21). Evidence suggests that CBD may have distinct neurobiological effects compared to THC (22, 23). Interestingly, preclinical data suggest that CBD appears to ameliorate psychotomimetic symptoms produced by THC (24), sensory gating deficits induced by NMDA receptor antagonists (25) and cannabinoid receptor agonists (26), and hyperlocomotion induced by amphetamine, cocaine, and ketamine, and stereotyped behavior induced by apomorphine in mice (27). Experimental studies in humans have demonstrated mixed findings regarding its antipsychotic properties of CBD (28-33). Cannabis that is high in THC and low in CBD is associated with a greater risk of psychosis (34-36) and some studies suggest that CBD may attenuate the psychotomimetic and anxiogenic effects of THC. Pretreatment with CBD has been observed to block the psychotomimetic effects of THC (26, 37, 38), although with mixed, dose-dependent effects (39). The literature on the behavioral, cognitive, and neurophysiological effects of THC, CBD, and their combination have been previously reviewed by us and others (5, 6). We briefly present this literature relevant to psychosis outcomes below.

In a small within-subject study, Bhattacharya et al (23) demonstrated that pretreatment with intravenous (IV) CBD (5 mg) attenuated the psychotomimetic effects of IV THC (1.25 mg). The absence of pharmacokinetic interactions suggested that the observed attenuation was possibly due to receptor-level interactions (23). In a between-subject, placebo-controlled study (n=48), Englund et al (40) examined the effects of oral CBD pretreatment (600 mg) on the psychotic symptoms induced by IV THC (1.5 mg) and found that a significantly lower proportion of subjects experienced clinically significant THC induced positive symptoms and episodic memory impairment with CBD pretreatment (40). Hindocha et al (41) examined the interaction of inhaled THC (8 mg) and CBD (16 mg) on human facial affect recognition in 48 volunteers. The authors noted that while the THC-CBD combination induced a subjective experience of feeling ‘stoned’ to the same extent as THC, the combination resulted in a significantly lower impairment in affect processing (41). Haney et al (42) examined the effects of pretreatment with four doses of oral CBD (0, 200, 400, 800 mg) on reinforcing, positive subjective, and physiological effects of smoked THC (5.3% - 5.8%) in 31 cannabis smokers and noted that none of the CBD doses altered any of the THC effects examined. More recently, Solowij et al (43) examined the effect of low (4 mg) and high (400 mg) doses of vaporized CBD on the intoxicating effects of vaporized THC (8 mg) in 36 frequent and infrequent cannabis users. Interestingly, the authors noted that the low dose of CBD potentiated the intoxicating effects of THC while the high dose of CBD attenuated the intoxicating effects of THC (43), suggesting that the proportion of THC and CBD in cannabis may influence subjective and behavioral effects.

Thus far, most of the investigations of THC and CBD interaction in human studies have focused on either behavioral or cognitive outcomes with only a few investigating neurophysiological outcomes. In a previous study from our group, we found no differences between THC and THC+CBD on P50 gating ratio (S2/S1) and evoked power (44). Preclinical studies on the mechanisms of the interaction between THC and CBD also report mixed results (45-50). The inherent challenges in modelling an acute behavioral response such as psychotomimetic effects in preclinical models after exposure to clinically translatable doses of THC and CBD further complicates the interpretability of these results.

In summary, the emerging data available thus far indicate that CBD may attenuate THC induced psychotomimetic effects while sparing the other subjective experiences. However, how CBD:THC ratio influences these interactions remains unstudied. Importantly, despite several reports comparing THC and CBD effects on psychophysiological outcomes, there are limited data examining the effects of CBD on THC induced psychosis relevant EEG outcomes. These interactive effects may also be sensitive to the dose and route of administration of THC and CBD. For instance, there is significantly greater variability in blood levels with the oral route (42) compared to IV (23).

In this study, we aimed to examine the effect of IV CBD on a range of IV THC induced subjective psychotomimetic and rewarding effects as well as a specific psychophysiological index of psychosis in healthy human participants as described below.

We have previously shown that neural noise, i.e., the randomness of signals, as captured by electroencephalography (EEG) signals with Lempel-Ziv complexity (LZC), is strongly related to the acute psychosis-like effects of THC (51). LZC is a nonlinear measure from information theory designed to assess the randomness (noise) of signals by counting the minimal number of distinct signal patterns (words) that are required to fully reconstruct the signal without information loss (52). LZC approaches zero in signals with regular and recurrent patterns (small number of distinct patterns) and approaches one as the randomness (large number of distinct patterns) of signals increases. In addition to predicting THC-induced psychotomimetic symptoms, increased LZC has been demonstrated in schizophrenia (53, 54) and specifically in patients in their early stages of illness with higher positive symptoms (55), potentially serving as an EEG biomarker of psychosis.

We hypothesized that CBD would attenuate THC-induced psychotomimetic effects and neural noise but not its rewarding effects. We further examined whether these interactions were dose related by testing three different doses of CBD (CBD:THC ratio 1:1, 2:1, 3:1). Lastly, we examined correlations between CBD’s effects on the subjective and electrophysiological psychosis-relevant effects of THC i.e. a) THC-induced psychotomimetic effects and b) THC-induced increase in neural noise.

## Results

### Sample Profile

The sample consisted of 28 (13 females) healthy volunteers with a median (IQR) age of 25.5 (10.25) years. The demographic and clinical details including the race, handedness, education, anthropometric measures, cannabis, and tobacco use patterns, schizotypal personality and psychosis proneness scores are presented in table 1. None of the subjects were cannabis naïve; 7 (25%) subjects reported cannabis use of two or more times per week. Of the 28 subjects who participated in phase-1, 10 (35.71 %) participated in phase-2 and received two additional doses of CBD:THC (1:1 and 3:1; supplementary figure 1). The demographic and clinical details are presented in table 1 and supplementary table 2. There were no statistically significant group differences between the 18 subjects who participated only in the phase-1 and the 10 subjects who participated in both phases of the study (supplementary table 2).

**Table 1:** Demographic variables.

| Variable | Summary – Phase 1<br>N = 28 | Summary – Phase 2<br>N = 10 (subset of 28) |
| --- | --- | --- |
| Age – median (IQR) | 25.5 (10.25) | 25 (10) |
| Gender – female - n (%) | 12 (42.86%) | 4 (40%) |
| Race – Caucasian: black: other (n) | 16:8:4 | 6:3:1 |
| Education – mean (SD) | 15.78 (2.41) | 15 (2.24) |
| Handedness – right – n (%) | 27 (96.42%) | 10 (100%) |
| Cannabis – frequent user – n (%) | 4 (14.28%) | 0 (0) |
| Smoker – n (%) | 5 (17.86%) | 2 (20%) |
| BMI – mean (SD) | 26.1 (4.8) | 25.9 (4.84) |
| NART – IQ – mean (SD) | 113.3 (5.63) | 111.8 (5.8) |
| Psychosis proneness - median (IQR) | 19.5 (17.5) | 20.5 (10.25) |
| SPQ score - median (IQR) | 3 (5) | 2.5 (6.75) |

### Behavioral, Subjective, and Physiological Measures

The outcomes of Positive and Negative Syndrome Scale (PANSS) positive, negative, general, and total scores; Clinician Administered Dissociative Symptom Sale (CADSS) - patient rated (PR) and clinician rated (CR) subscale scores; and subjective experiences measured with Visual Analog Scale (VAS) – “VAS high” and “VAS anxious”, scores in phase-1 and phase-2 of the study are listed in table 2.

**Table 2:** Behavioral, physiological and neural noise outcome measures.

| Variable | Phase 1 |  | Phase 2 |  |  |  |
| --- | --- | --- | --- | --- | --- | --- |
|  | Main effect of drug condition - | Post-hoc (THC vs CBD 5 mg) | Main effect of drug condition | Post-hoc (THC vs CBD 2.5 mg) | Post-hoc (THC vs CBD 5 mg) | Post-hoc (THC vs CBD 7.5 mg) |
| PANSS positive ATS (p value) | 16.01 (<0.00001) | 0.58 (0.44) | 8.73 (<0.00001) | 7.83 (0.005) * | 1.696 (0.19) | 1.25 (0.26) |
| PANSS positive responder # ATS (p value) | 21.2 (<0.00001) | 9.73 (0.0018) * | NA | NA | NA | NA |
| PANSS negative ATS (p value) | 8.93 (0.00005) | 0.84 (0.35) | 4.91 (0.002) | 1.35 (0.24) | 3.38 (0.066) | 0.36 (0.55) |
| PANSS general ATS (p value) | 26.74, (0.00001) | 3.4 (0.065) | 18.08 (<0.00001) | 5.16 (0.023) | 1.41 (0.24) | 0.08 (0.78) |
| PANSS total ATS (p value) | 25.82 (<0.00001) | 1.6 (0.21) | 17.58 (<0.00001) | 11.11 (0.0009) * | 1.48 (0.22) | 0.002 (0.96) |
| PANSS total responder # ATS (p value) | 18.64 (<0.00001) | 6.78 (0.0092) * | NA | NA | NA | NA |
| CADSS – PR ATS (p value) | 17.25 (<0.00001) | 0.07 (0.79) | 4.48 (0.006) | 0.67 (0.41) | 1.12 (0.29) | 0.21 (0.65) |
| CADSS – CR ATS (p value) | 29.98 (<0.00001) | 1.93 (0.16) | 11.11 (<0.00001) | 3.03 (0.08) | 0.91 (0.34) | 3.02 (0.08) |
| VAS – high | 43.02 (<0.00001) | 0.13 (0.72) | 17.57 (<0.00001) | 0.15 (0.71) | 0.11 (0.73) | 0.15 (0.698) |
| ATS (p value) |  |  |  |  |  |  |
| VAS – anxiety<br>ATS (p value) | 7.24 (0.0002) | 0.08 (0.78) | 2.69 (0.04) | 4.01 (0.04) | 0.68 (0.41) | 10.41 (0.001) * |
| Pulse rate<br>ATS (p value) | 37.9 (<0.00001) | 2.44 (0.12) | 8.85 (<0.00001) | 0.09 (0.76) | 2.53 (0.11) | 0.08 (0.78) |
| Neural noise (LZC)<br>ATS (p value) | 5.66 (0.0015) | 0 (NA) | 2.68 (0.04) | 8.83 (0.003) * | 0.33 (0.56) | 0.76 (0.38) |

### THC-Induced Psychotomimetic Effects – PANSS Positive Symptom Scores (Table 2)

#### Phase -1

There was a significant main effect of treatment on PANSS positive change score (Anova Type Statistic [ATS] = 16.01, df = 2.87, p < 0.00001). Compared to THC, CBD:THC-2:1 resulted in a decreased PANSS positive score, that was not statistically significant in a pairwise post-hoc comparison (ATS = 0.58, df = 1, p = 0.44) (figure 1a). Thirteen of the 28 subjects had a clinically meaningful psychotomimetic response to THC. In this ‘responder’ subgroup, the CBD:THC-2:1 had a significantly lower PANSS positive change score compared to THC alone (ATS = 9.73, df = 1, p = 0.0018) (figure 1b).

**Figure 1:**
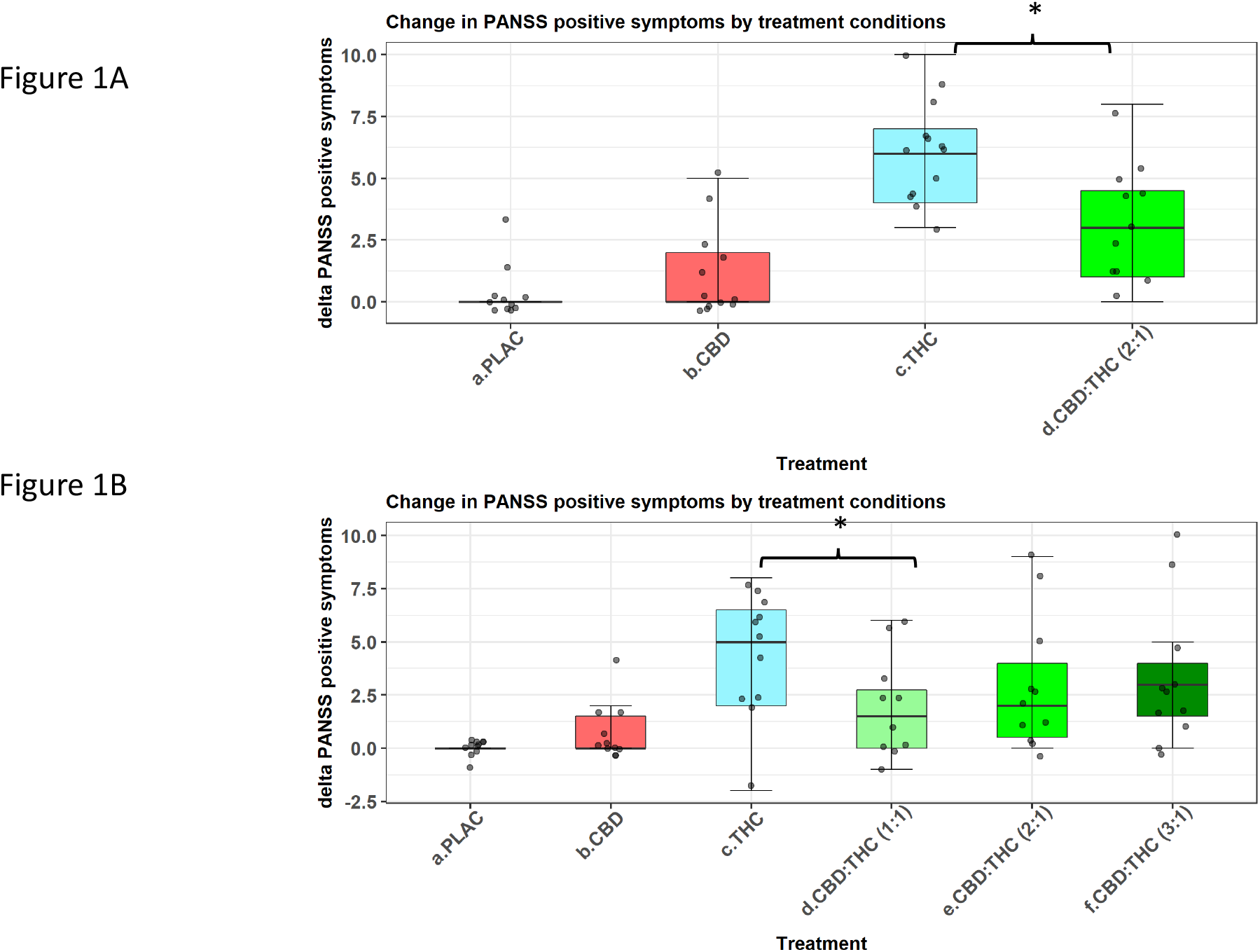
Box and jitter plots showing the change in PANSS positive symptoms from baseline the 15 minutes post infusion across the 4 treatment conditions. 1A. Phase-1 responders – a statistically significant group difference is noted between THC and CBD:THC(2:1) conditions; 1B. Phase-2 – a statistically significant group difference is noted between THC and CBD:THC(1:1) conditions but not the 2:1 and the 3:1 combinations.

#### Phase -2

In the analysis of the phase-2 (figure 1c) comparing THC, CBD:THC-1:1, CBD:THC-2:1, and CBD:THC-3:1, there was a significant main effect of the treatment on the PANSS positive change score (ATS = 8.73, df = 3.37, p < 0.00001). In post-hoc analysis involving pair-wise comparisons between THC and the three CBD:THC combinations, the lowest THC-induced PANSS positive score was noted with CBD:THC-1:1 (ATS = 7.83, df = 1, p_corr_ = 0.015). The CBD:THC 2:1 and 3:1 doses also resulted in lower PANSS positive scores compared to the THC alone, but the differences were not statistically significant. Overall, there was an inverse dose response relationship between the CBD dose in the CBD:THC combinations and the magnitude of attenuation of THC-induced positive symptoms (figure 1c).

There were no differences between THC and CBD:THC conditions in the PANSS negative or general symptom scores (supplementary figure 2 and 3). Although not statistically significant, consistent with the above results, THC-induced PANSS general symptom score was also maximally attenuated in the CBD:THC-1:1 condition (ATS = 5.15, df = 1, p_corr_ = 0.069). For PANSS total score, phase -1 responder analyses revealed significantly lower scores with CBD:THC-2:1 condition compared to THC (ATS = 6.78, df = 1, p = 0.009). In phase-2, PANSS total score was significantly attenuated only in the CBD:THC-1:1 condition (ATS = 11.11, df = 1, p_corr_ = 0.002) (table 2; supplementary figure 4).

### Perceptual Alterations – CADSS - Patient Rated and Clinician Rated scales (Table 2)

#### Phase-1

There was no significant main effect on CBD treatment on THC induced perceptual alterations. CBD:THC-2:1 resulted in a marginal reduction in the THC CADSS PR and CR scales, but the differences were not statistically significant.

#### Phase-2

None of the three CBD:THC combinations were significantly different from the THC condition on CADSS PR or CR. However, the largest relative reduction in THC-induced perceptual alteration measured by CADSS CR was noted in CBD:THC-1:1 condition when compared to the 2:1 and 3:1 ratios, consistent with the pattern observed on PANSS positive scores (table 2, supplementary figures 6, 7).

### Subjective Effects – VAS “High” and “Anxious” (Figures 2a and 2b)

**Figure 2:**
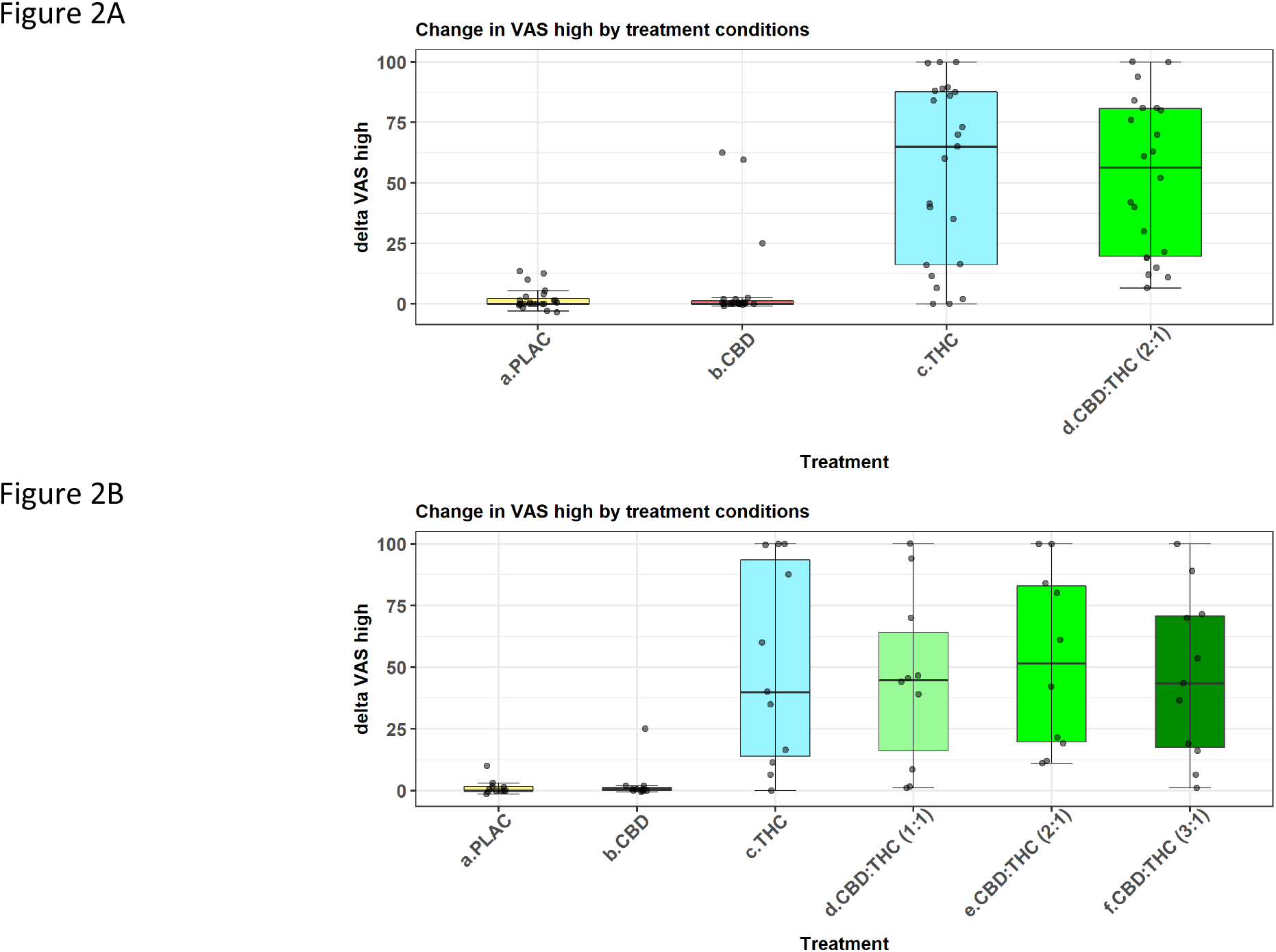
Box and jitter plots showing the change in subjective effects of high measured on Visual Analog Scale from baseline the 15 minutes post infusion across the 4 treatment conditions. 2A. Phase-1 – No significant group difference is noted between THC and CBD:THC(2:1) conditions; 2B. Phase-2 – No significant group differences are noted between THC and CBD:THC(1:1, 2:1 and 3:1) combinations.

#### “High”

*<u>In phase-1</u>*, there was a main effect of the treatment on subjective experience of high (ATS = 43.03, df = 2.24, p <0.00001), but there was no significant difference between the THC and the CBD:THC-2:1 in the post-hoc analysis (ATS = 0.13, df = 1, p = 0.72).

*<u>In phase-2</u>*, there was a main effect of treatment (ATS 17.57, df = 2.97, p < 0.00001) on “High”. However, there was no difference between the THC and the three CBD:THC combinations – CBD:THC-1:1 (ATS = 2.04, df = 1, p_corr_ = 0.45), 2:1 (ATS = 0.11, df = 1, p_corr_ > 0.99) and 3:1 ratios (ATS = 3.96, df = 1, p_corr_ = 0.14).

#### “Anxiety”

*<u>In phase-1</u>*, there was a main effect of treatment but no significant difference between THC and CBD:THC-2:1 conditions (table 2). However, in *<u>phase-2</u>*, THC-induced anxiety was maximally attenuated with the combination of CBD:THC-3:1 and this difference was statistically significant (ATS = 10.41, df = 1, p_corr_ = 0.004) (supplementary figure 5).

### Physiological Effects

In *<u>phase-1</u>* there was a main effect of treatment (ATS = 37.9, df = 2.5, p = <0.00001) and both THC and CBD:THC-2:1 resulted in a significant increase in heart rate at +15 minutes. However, there was no difference between THC and CBD:THC-2:1 conditions in the post hoc analysis (ATS = 2.44, df = 1, p = 0.12). Similarly, in the *<u>phase-2</u>*, all treatment conditions resulted in a robust increase in heart rate compare to placebo but there was no statistically significant difference between THC and CBD:THC conditions (supplementary figure 8).

### Neural Noise (Table 2 and Figures 3a and 3b)

**Figure 3:**
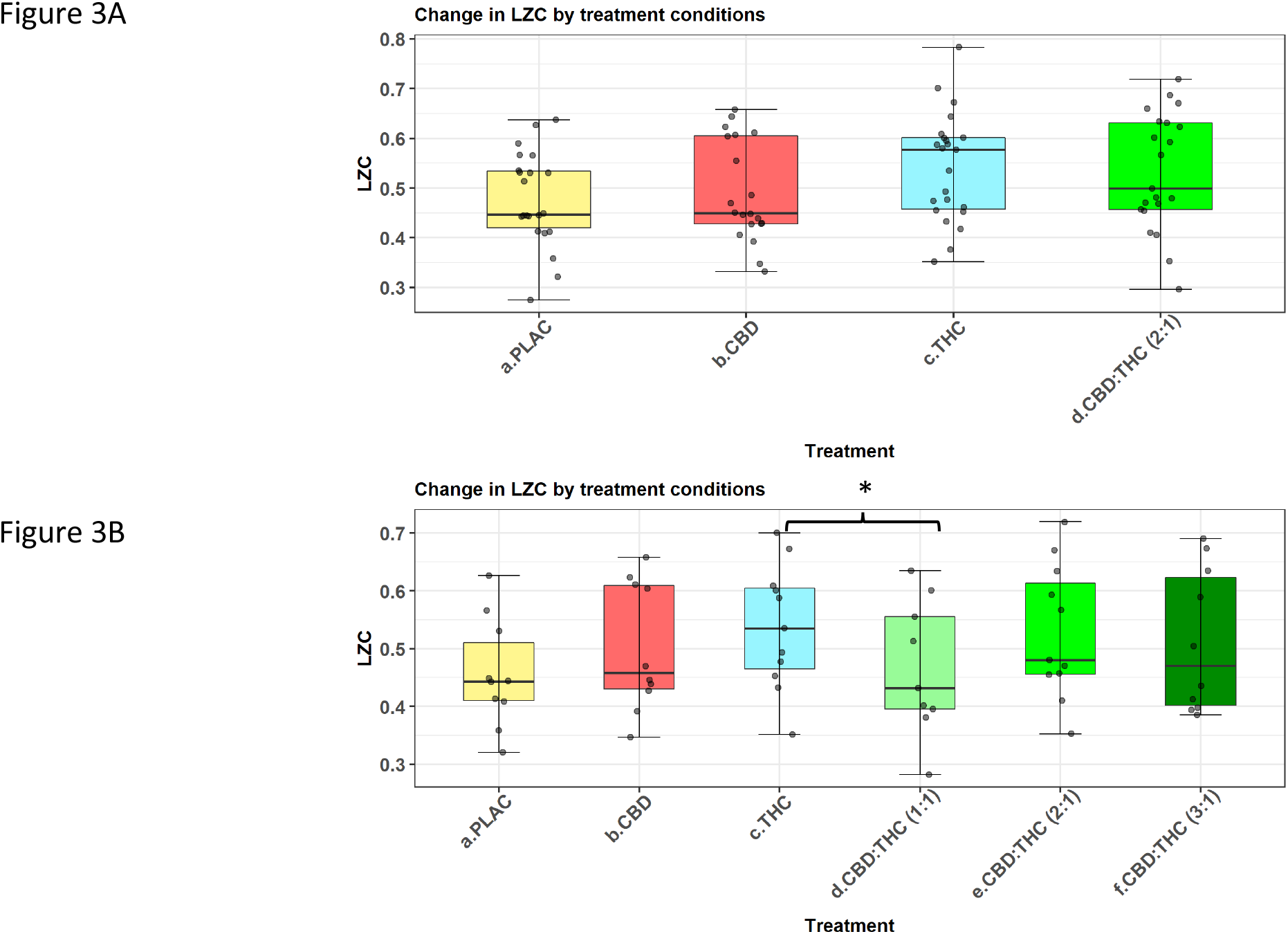
Box and jitter plots showing post infusion from neural noise measured as Limpelziv complexity across the 4 treatment conditions. 3A. Phase-1 – No significant group difference is noted between THC and CBD:THC(2:1) conditions. A similar pattern was noted in phase 1 responders (not shown); 1B. Phase-2 – a statistically significant group difference is noted between THC and CBD:THC(1:1) conditions but not the 2:1 and the 3:1 combinations.

#### Phase-1

There was a main effect of treatment on LZC with an increase in LZC noted in both the THC and the CBD:THC-2:1 conditions compared to placebo (ATS = 5.66, df = 2.52, p = 0.0015). Post-hoc analysis revealed that the CBD:THC-2:1 condition had a lower median LZC compared to THC alone, although the differences were not statistically significant (ATS = 0, df = 1, p = >.05) (figure 3A). A similar pattern was observed among THC PANSS responders, where we noted a modest but statistically non-significant reduction in the reduction in LZC (ATS = 0.2, df = 1, p > 0.05).

#### Phase-2

There was a main effect of treatment (ATS = 2.68, df = 3.3, p = 0.04) and all three CBD:THC combinations resulted in a lower median LZC compared to THC alone (figure 3B). Post-hoc analysis revealed that the maximal attenuation of THC-induced increase in LZC was in the CBD:THC-1:1 condition and this difference was statistically significant (ATS = 8.83, df = 1, p_corr_ = 0.009), consistent with the pattern observed with the THC induced PANSS positive scores.

### Relationship Between CBD Attenuation of Positive Symptoms and Attenuation of Neural Noise

Correlations between changes in PANSS positive and total scores and changes in LZC were examined in the phase-1 subjects and phase-1 responders. Small statistically non-significant positive correlations were noted between change in LZC and change in positive symptoms from placebo to THC condition in phase-1 (r = 0.13, p > 0.05) and in the subsample of phase-1 responders (r = 0.28, p > 0.05). Among the responders a large, marginally significant correlation was noted between change in LZC and change in the PANSS total symptoms (r = 0.64, p = 0.07). There were no correlations between change in positive or total symptom scores with change in LZC from THC to CBD:THC conditions in Phase-1 or Phase-1 responders.

## Discussion

We present novel results demonstrating dose dependent interactions between IV CBD and IV THC on subjective and objective, psychosis and reward related measures, in this double-blind, randomized, placebo-controlled two-phase human laboratory study in healthy individuals. Pretreatment with IV CBD 5mg (∼ 2:1 CBD: THC ratio) attenuated THC induced psychotomimetic symptoms without affecting any other subjective, behavioral or cognitive effects. This attenuation was particularly notable in those ‘responders’ who developed a clinically meaningful psychotomimetic response to THC alone (≥ 3 increase in PANSS positive symptoms). Furthermore, we found that the maximal attenuation of THC induced psychotomimetic symptoms resulted from the lowest tested dose of CBD (2.5 mg) (∼ 1:1 CBD: THC ratio) and an inverse linear dose-response relationship was observed between CBD dose and attenuation of THC induced psychotomimetic response.

Consistent with the effects on THC induced psychotomimetic response, CBD treatment was associated with a decrease in THC induced neural noise, a biomarker of drug induced psychotomimetic effects (51). We observed that the largest decrease in neural noise was also with the lowest dose of CBD (2.5 mg) (∼ 1:1 CBD: THC ratio), similar to the psychotomimetic response. However, there was no correlation between the decreases in the neural noise and subjective psychotomimetic effects of THC. Finally, in contrast to psychotomimetic symptoms, it is noteworthy that none of the doses of CBD decreased the rewarding effects of THC.

The results of this study provide novel insights into the intricate psychopharmacology of cannabinoids important for future studies on cannabis and cannabinoid psychopharmacology as discussed below.

### CBD and THC Interactive Behavioral Effects

The attenuation of THC induced psychotomimetic effects by CBD pretreatment, is in agreement with the findings of two earlier studies (Bhattacharya et al (23), and Englund et al (40)). Both studies used PANSS positive symptom subscale to measure THC-induced psychotomimetic effects similar to this study. Bhattacharya et al examined the interactive effects of single doses of IV THC (1.25mg) and IV CBD (5 mg), while Englund et al. tested 600 mg of oral CBD 210 minutes prior to IV THC (1.5 mg). Given the oral bioavailability of CBD is ∼6% (56), the dose in Englund et al, would be equivalent to ∼3.6 mg of IV CBD although, additional variability of the peak plasma concentration and half-life is recognized with the oral route (21, 56). Thus, both these studies examined higher CBD:THC ratios (∼2.4-4:1) compared to ∼2:1 in phase-1 of our study and 1:1 and 3:1 in Phase -2. Further, it was the lowest CBD:THC ratio (1:1) examined in phase-2, associated with the maximal attenuation of THC-induced psychotomimetic effects with an inverse linear dose response relationship between CBD and THC induced psychotomimetic effects. These data would suggest that equivalent ratio (1:1) of CBD:THC in herbal cannabis may result in the least psychotomimetic effects. However, since we did not examine lower CBD:THC ratio < 1 in this study, it is not possible to estimate the impact of lower CBD concentrations in most current herbal cannabis.

In contrast to the psychotomimetic response, the experience of subjective effect of high was not significantly different in any of the three CBD:THC dose ratios. This relative absence of effects of CBD on ‘high’ and other related subjective effects has been demonstrated previously by Hindocha et al (41) and Haney et al (42). Using inhalational administration of CBD 16 mg and THC 8 mg (CBD: THC 2:1), Hindocha et al, noted that the subjective feeling of being ‘stoned’ was not altered by the combination of CBD and THC compared to THC alone. Similarly, Haney et al, noted that oral CBD (200, 400 and 800 mg) administered 90 minutes prior to smoked cannabis (5.3-5.8% THC), did not alter the subjective effects of ‘high’ and ‘good effect’ associated with THC. In contrast, Solowij et al (43), reported that low dose vaporized CBD (4 mg) *augmented* the intoxicating effects of vaporized THC (8 mg) while a higher dose of vaporized CBD (400 mg) resulted in an attenuation of the intoxicating effects measured with CADSS clinician-rated subscale. These effects were specifically prominent in the infrequent cannabis users. In contrast, in our analyses none of the CBD:THC combinations were significantly different on the CADSS subscale scores. It is of interest that Solowij et al observed an augmentation of intoxication at a CBD:THC ratio of 1: 2 (THC dominant) and attenuation of intoxication at a ratio of 50:1-both ratios that we did not test, but suggest that a wide dose range of CBD:THC should be further studied. Lastly, we noted a statistically significant difference between the THC and CBD:THC (3:1) condition on subjective effects of anxiety. Although, both 2.5 mg and 7.5 mg produced greater anxiolysis compared to the 5 mg condition in the phase-2 analysis (Supplementary figure 5)-this was statistically significant only with the higher CBD dose. This is consistent with the inverted U-shaped dose-response of CBD on anxiety that has been previously reported by Linares et al, using 150 mg, 300 mg and 600 mg of oral cannabidiol in a simulated public speaking test (57).

### CBD and THC and Neural Noise

To our knowledge, this is the first study to examine neural noise (LZC), an electrophysiological correlate of psychosis, to assess the effects of THC, CBD, and their interaction. Neural noise has been shown to be elevated in acute phases of schizophrenia (54, 55) and we have previously demonstrated it to be strongly correlated with THC-induced psychotomimetic effects, specifically with positive- and disorganization-like symptoms (51). We now demonstrate that CBD-related attenuation of the psychotomimetic effects of THC was mirrored by a similar attenuation of THC-induced noise. Further, examination of dose-related effects demonstrates a trend towards an inverted U-shaped response as CBD 2.5 mg and CBD 7.5 mg resulted in a larger attenuation of neural noise compared to CBD 5 mg. Future studies in larger samples with a wider range of dose ratios of CBD:THC will be needed to delineate this nonlinear dose-response relationship. CBD treatment has been associated with improvement in psychotic symptoms in patients with psychotic disorders in some clinical trials but not others (58-61). Whether CBD treatment has an effect on the elevated neural noise observed in acute schizophrenia, has not been reported thus far. Future studies that integrate psychophysiological measures of neural noise may shed further light on the potential neural mechanisms by which CBD exerts its antipsychotic effects in specific subpopulations of psychosis.

### Possible Mechanisms of CBD Antagonism of THC-Induced Psychotomimetic Effects

Several pre-clinical and clinical studies have demonstrated opposing effect of CBD on specific THC-induced behavioral, cognitive, and biological outcomes. The opposing effects of THC and CBD on human cognition has been extensively reviewed (5, 6, 62) and the opposing effects on specific cognitive domains are possibly mediated via differential effects of these compounds on functional connectivity between prefrontal cortex, hippocampus, and dorsal striatum (63). We discuss findings from preclinical, molecular, human imaging studies relevant to the opposing effects of CBD on THC-induced psychotomimetic effects. In rodents, co-administration of CBD has been demonstrated to attenuate acute THC-induced c-Fos expression in multiple brain regions (48). c-Fos is one of the immediate early genes with altered expression during learning and memory and has also been proposed to be a biomarker of schizophrenia (64, 65). More relevant to the timescale of the psychotomimetic response to THC, acute intra-PFC infusion of CBD has been shown to reverse the cognitive impairments caused by glutamatergic antagonism suggesting that the therapeutic effects of CBD may be relevant only during pathologically aberrant states (47). In contrast, striatal glutamate increase has been shown in human studies to be a marker of THC-induced acute psychotomimetic response (66). More recently, a regional specificity in THC-CBD interaction relevant to alleviation of psychotomimetic response has been demonstrated by Wall et al (67). Using resting state functional magnetic resonance imaging, the authors noted that CBD:THC combination resulted in alleviation of THC induced functional connectivity disruption, specifically in the limbic sub-division of the striatum but not in the associative and sensorimotor sub-striatal networks (67). Taken together, the findings suggest that restoring cortical-striatal balance of glutamatergic tone may be a potential underlying mechanism of CBD action in attenuating THC-induced acute psychotomimetic response. CBD has been demonstrated to act via various receptor and enzymatic mechanisms in several biological pathways. In addition to CB1 receptors, CBD has been shown to have effects on the Adenosine receptor A_2A_ and 5 hydroxy tryptamine receptor 1A (5HT-1A), GPR 55, and several receptors of the TRPV family in the brain (68-70). However, the antagonism of THC induced psychotomimetic effects by CBD is possibly mediated through negative allosteric modulation of

CB1 receptor (70). The precise mechanisms of THC and CBD interactions at the receptor level in the brain still needs to be characterized in future studies that employ both human receptor imaging studies as well as preclinical studies that permit in vivo assay of molecular and neurochemical alterations under simultaneous drug exposure conditions.

### Clinical and Public Health Relevance of the Findings

The results of our human laboratory study align with and confirm some of the previous findings in epidemiological studies on the effect of CBD content in cannabis on psychotic experience in the community. Morgan and Curran analyzed hair samples in 140 individuals and noted higher schizophrenia-like positive symptoms in persons with THC-alone detected in hair samples compared to persons with both THC and CBD (71). In another study, Morgan et al, demonstrated that compared to low CBD:THC (∼ 1:100) cannabis users, high CBD:THC (∼1:3) cannabis users had reduced attentional bias towards food and drug stimuli. However, there were no differences between the groups in the subjective feeling of being ‘stoned’ (72). The authors suggested that relatively higher CBD:THC ratio may a have protective effect against developing cannabis dependence. Schubart et al have also reported an inverse relationship between CBD content and the severity of positive psychotic experiences in a large Dutch community study. Interestingly, highest CBD concentration that the authors noted in their sample was CBD:THC of 1:2 and the lowest being 1: 81.5 (73). Taken together, the results of our investigation and these naturalistic studies suggest that relatively higher CBD:THC ratio up to 1:1 may optimally protect against psychotomimetic and dependence forming effects while minimally affecting the subjective affective experiences of THC in cannabis. This hypothesis needs further investigation and has the potential to inform harm reduction strategies and regulations for marketing of cannabis with specific CBD:THC ratios especially given the increasing legalization of recreational cannabis and medicinal cannabinoids in the US and the rest of the world. The attenuation of psychotomimetic effects of THC at 1:1 ratio of CBD:THC is of particular interest as this is the ratio of CBD:THC in the currently approved nabiximol preparations for the treatment of spasticity in Multiple Sclerosis (74, 75).

A few limitations should be considered while interpreting the results of our study. The dose related effects of CBD (phase 2) were tested in a small sub-sample of subjects. Although we observed a large and significant effect for CBD attenuation of THC psychotomimetic symptoms and neural noise at the lowest dose (1:1) tested, this needs to be replicated in a larger study perhaps with a wider dose range CBD:THC ratios (for instance ratios <1). Although consistent with some population-based studies, the generalizability of these results to the effects of cannabis exposure in the real-world setting remains complicated by the potential moderating effect of other phyto-cannabinoids and chemical compounds in herbal cannabis. Further, IV administration of THC and CBD, chosen for more precise and reliable delivery of these compounds limits the generalizability to the typical smoked or oral routes of cannabis consumption. This study focused on specific psychosis related behavioral and psychophysiological interactions of CBD and THC. A recent study has demonstrated a significant impairment in driving performance up to 2 hours after administration of CBD:THC (1:1) cannabis (76). Thus, despite potential benefits of 1:1 CBD:THC ratio on psychosis outcomes, a wider range of motor and real-life performance effects warrants further study. Finally, future studies need to examine repeated or chronic dosing effects of CBD and THC combinations to fully understand the range of interactions in humans.

## Conclusions

CBD attenuates THC-induced psychotomimetic symptoms in healthy humans and the effect is more notable in those who develop a robust psychotomimetic response with THC. Further, amongst the CBD:THC dose ratios studied, a 1:1 ratio produces maximal attenuation of psychotomimetic effects compared to 2:1 and 3:1 with an inverse linear dose response. The attenuation of psychotomimetic symptoms is mirrored by a similar attenuation of THC-induced increase in neural noise, an electrophysiological biomarker of psychosis. Lastly, none of the CBD doses tested resulted in attenuation of the THC-induced subjective experience of ‘high’. Future studies in larger samples with a wider range of dose ratios and longer dosing paradigms are needed to systematically examine the spectrum of interactive effects between CBD and THC.

## Online methods

### Study Design

This was a two-phase, double-blind, randomized, counterbalanced, placebo-controlled cross-over trial. In phase-1, all subjects participated in the four test days where they received one of 4 drug conditions (Placebo, THC [0.035mg/Kg; ∼2.5mg in a 70kg individual], CBD 5mg, THC ∼2.5mg + CBD 5 mg) on each test day separated by at least 3 days. Subjective, behavioral, physiological, and EEG data were collected before and at several time points after drug administration on each test day. In phase-2, a subsample of subjects from phase-1 participated in two additional test-days to specifically examine the interactions of THC with two additional doses of CBD (THC ∼2.5mg + CBD 2.5 mg, THC ∼2.5mg + CBD 7.5 mg). A representation of the study design is presented in supplementary figure 1. The study protocol was approved by the Human Investigations Committee at the Yale University and the Human Subjects Study committee of the VA Connecticut Healthcare System (VACHS). The study was carried out at the Neurobiological studies unit of the VACHS under an existing IND for the administration of THC (#51671) and CBD (#101185).

### Participants

The subjects consisted of 28 (13 females) healthy volunteers recruited using methods consistent with our previous HLS (77, 78). After obtaining written informed consent, subjects underwent a detailed medical and psychiatric evaluation, laboratory tests, electrocardiogram, and urine toxicology. Structured Clinical Interview (SCID) for DSM-IV (non-patient) for healthy controls was conducted to exclude psychiatric diagnoses. General intelligence was assessed with National Adult Reading Test. Subjects were asked to explicitly consent to identifying a collateral informant for verification of inclusion/exclusion criteria. Cannabis naïve individuals were excluded from the study to avoid the theoretical risk of de novo exposure in the laboratory. Similarly, subjects with major DSM IV axis 1 disorders were excluded to avoid the potential risk of illness exacerbation.

### Drugs - THC Dose, Rate, and Route of Administration

The route, dose, and rate of administration of THC influence its behavioral and cognitive effects (78). The THC doses used in our studies (78) produced blood levels comparable to those achieved by recreational cannabis use and mimic the time course of plasma THC levels associated with the clinical “high” from smoking 0.5-1.5 of a standard NIDA cannabis cigarette (79-81). Even with standardized-pace smoking procedures, subjects are still able to titrate the dose of cannabinoids they receive and in doing so, negate attempts to deliver a uniform dose (82). The IV route of administration was chosen to reduce inter and intra-individual variability in plasma THC levels with the inhaled route and to mimic the time course of plasma THC levels associated with the clinical “high” (78). The dose chosen for this study was based on several previous studies conducted by our group and has been demonstrated to be well tolerated by subjects while producing transient but reliable psychotomimetic effects (83). THC [0.035 mg/kg (∼ 2.5 mg in a 70 kg individual)], was administered IV over 20 minutes into a rapidly flowing infusion of normal saline. Similar doses in previous studies have produced clinically and statistically significant behavioral and cognitive effects. Placebo administration involved an equal volume - 2ml of 190 proof ethanol.

### CBD Dose, Rate, and Route of Administration

In experimental studies, CBD has been safely administered to healthy humans in doses ranging from 15 mg to 600mg (oral) per day (20, 84-97) alone and in combination with THC and is known to have been tolerated well with no significant adverse effects. Previous studies have also demonstrated that IV CBD is well tolerated at the dose ranges used in this study (2.5 – 7.5 mg), and have produced effects similar to oral CBD (23). While most studies have administered CBD orally, the low and variable oral bioavailability of CBD presents a limitation. For instance, the oral absorption of CBD of 600 mg produces plasma levels of 4.7 (SD=7) and 17 (SD=29) ng/ml at 1 and 2 hours post-administration, respectively (22). Thus, in this study 3 active doses of IV CBD were administered, on separate test days. In an average weighing individual, **the three doses of CBD, 2.5mg, 5mg, and 7.5mg IV**, resulted in CBD:THC ratios equivalent to 1:1, 2:1, and 3:1, respectively. Additional details regarding the source, preparation, storage, stability, and quantitative analysis of cannabidiol doses are presented in the supplementary methods.

### Test-Day Procedures

An overview of the schedule of test-days is presented in supplementary table 1. Subjects were instructed to fast overnight and to report to the test facility at 8 am. Subjects were not permitted to drive to and from the test facility. Exclusions (like urine drug and pregnancy test) were confirmed on each test morning. IV lines were inserted, and baseline behavioral, subjective, physiological, and neurochemical assessments described below were collected. CBD (active or placebo) was administered over 2 minutes immediately followed by THC (active or placebo) through an IV line over 20 minutes. Outcome measures were repeated periodically as described below and enumerated in supplementary table 1.

### Behavioral and Subjective Measures

#### Psychosis and perceptual alterations

Positive, negative, and general symptoms were assessed using the positive, negative, and general symptoms subscales of the Positive and Negative Syndrome Scale (PANSS) (98). Perceptual alterations were measured using the Clinician Administered Dissociative Symptoms Scale (CADSS) (99). The CADSS is a scale consisting of 19 self-report items and 8 clinician-rated items (0 = not at all, 4 = extremely) that we have previously shown to be sensitive to the psychotomimetic effects of THC (78) (fig. 1). The scale captures alterations in environmental/time/body perception, feelings of unreality, and memory impairment.

#### >Subjective Effects of Cannabis

Feeling states associated with cannabis intoxication were measured using a self-reported visual analog scale of four feeling states (“high” and “anxious”) associated with cannabis effects (99-101). Subjects were asked to score the perceived intensity of their current feeling states by making a mark on a 100 mm line printed on a paper sheet (0 mm from the line’s left end = ‘not at all’; 100 mm from the left = ‘extremely’). These data were captured to validate that the experiment is relevant to cannabis effects and to determine how CBD pretreatment alters the subjective effects of THC.

#### Long-term follow-up of cannabis use

Subjects were evaluated 1 and 3 months after study completion to document whether study participation altered future cannabis consumption (78).

### EEG Paradigm and Data Acquisition

Electroencephalography (EEG) data were recorded at the scalp using a 64-channel (extended 10-20 system) ActiveTwo Biosemi system (Biosemi B.V., Amsterdam, Netherlands) at a sampling rate of 1024 Hz with an on-line low-pass filter of 256 Hz to prevent aliasing of high frequencies. All electrodes were referenced during recording to a common-mode signal (CMS) electrode between and then subsequently re-referenced to the nose offline.

During recordings, subjects were seated in a comfortable chair in front of a monitor in a darkened, sound-attenuated, electromagnetically shielded booth while listening to auditory stimuli were delivered through via insert earphones (Etymotic Research, Inc., Elk Grove Village, IL, USA) as part of an auditory oddball task described elsewhere (102, 103). Briefly, the task consisted of a series of frequent (83.33%) ‘standard’ tones, infrequent (8.33%) ‘target’ tones, and infrequent (8.33%) task-irrelevant ‘novel’ distractor sounds presented with a 1250 ms stimulus onset asynchrony in three separate blocks of 180 stimuli each. Standard tones were 500 ms in duration, target tones were 500 ms in duration, and novel distractor sounds ranged between 175 and 250 ms in duration. All stimuli were presented binaurally using an intensity of 80 dB SPL (C weighting). The task was presented using Presentation software (Neurobehavioral Systems, Berkeley, CA, USA). EEG collection was continuously monitored on a screen outside the recording booth to detect drowsiness. Only EEG data preceding each stimulus were used for neural noise (LZC) estimation.

### EEG Preprocessing

In order to minimize the confounding effects that artifactual contamination could have on measuring neural noise in EEG signals, raw data were cleaned and only Cz was used for analyses, given that this electrode tends to be the least contaminated by artifactual noise resulting from muscle activity (104). Raw EEG data were preprocessed following standard procedures as described elsewhere (51, 103). Briefly, EEG data were band-pass filtered (0.5-100Hz) with a windowed sinc FIR filter (−6dB/Hz), 60Hz line noise was removed using a multi-tapering technique (105), muscle artifacts were cleaned using a canonical correlation analysis-based blind source separation technique (106), and eye movement and blink artifacts were removed with an adaptive filtering technique (107). Data were segmented in 1250ms epochs time-locked to stimulation onset, with a 250ms pre-stimulus baseline. A ±95 μV criterion was used to remove bad trials.

### Neural Noise (LZC) Computation

Following previous work from our group on the psychotic-like effects of THC (51), we calculated neural noise (LZC) on the pre-stimulus (−250s to -1ms) preparatory baseline period of each trial of an P300 oddball task. In that study, we showed that LZC during each trial’s the expectation/focused attention period was strongly related to the psychotomimetic effects of THC (51). The calculation of LZC of a real-valued signal requires to encode it into a symbolic sequence. Following previous work (51), we used the median-crossing approach which is robust against outliers to binarize the pre-stimulus segment (−250ms to -1ms) of each trial’s EEG signal at Cz. LZC was calculated using Lempel and Ziv’s exhaustive parsing algorithm (52) and then normalized between 0 and 1. Each subject’s LZC value was obtained by averaging LZC across trials. EEG analyses were conducted using Matlab 2020b (MathWorks, Natick, MA,USA) custom scripts and the EEGLab toolbox (108).

### Statistical Analyses

The distribution of each outcome variable was visually examined for normality with histogram, box, and whisker plot and normality was tested using Kolmogorov-Smirnov statistics. The behavioral and subjective data (PANSS, CADSS, VAS feeling states) were examined as the change from baseline at post-infusion 15 minutes (P15) time-point, as both the physiological and psychological effects of THC have been shown to peak around this time in IV infusion studies (78). This time-point was also closest to the post-infusion EEG measure at +25 minutes. The PANSS positive symptoms, VAS high, and EEG neural noise (LZC) were tested as the primary outcomes and the remaining PANSS measures, CADSS patient and clinician rated scores, VAS scores and physiological measures of outcome (pulse, blood pressure) were examined as secondary outcomes. The effects of treatment condition on the outcome measures were tested using linear mixed model with treatment as a within subject fixed factor and subject ID included in the model as a random variable. For the analysis of phase-1 data, the drug-condition factor involved four levels (Placebo, THC, CBD, CBD:THC-2:1) and in phase-2 it involved testing two additional levels in a subsample of participants (CBD:THC-1:1 CBD:THC-3:1). As previously described (40, 109), we examined the CBD attenuation of positive symptoms in a subsample of “THC-responders”, i.e. those who had a clinically significant psychotomimetic response to THC, defined as a change in PANSS positive score ≥ +3 with THC alone in phase-1. In the event of non-normal distribution of outcome residuals or a singular model fit due to invariant variance-covariance structure, non-parametric model for the analysis of factorial experiments (Brunner et al, 2002) (110) was used to examine the drug-condition effects. Both these models offer the advantage of greater flexibility in fitting correlated factor structure in repeated measures and are optimal when data are missing at random. Further, the latter non-parametric models are also robust for smaller sample sizes (111-113). Post-hoc comparisons were defined *a priori* for the comparison between THC and CBD:THC-2:1 condition in the analysis of phase-1 data and between THC and the three different CBD doses in the CBD:THC combinations for the analysis of phase-2 data. Bonferroni corrections were applied for multiple comparisons within outcome domains. The relationship between CBD attenuation of THC-induced positive symptoms and neural noise was visually examined with scatter plots and the relationship was measured using Pearson’s correlation coefficient. Data were plotted and analyzed in R version 4.0.1 (R: A language and environment for statistical computing) using packages Tidyverse (114), lme4 (115), lmerTest (116), and nparLD (113).

## Data Availability

Data will not be made available at this stage of pre-print publication

