## supplementary table for "Delta-9-tetrahydrocannabinol and cannabidiol in psychosis: A balancing act of the principal phyto-cannabinoids on human brain and behavior?"

**Preparation of IV cannabidiol**

**Source:** Cannabidiol was manufactured and supplied by STI pharmaceuticals Ltd. (IND#101185).

**Preparation:** Cannabidiol is noted to be soluble in ethanol up to 36mg/ml as reported by our supplier and noted by an independent source of Cannabidiol, Cayman Chemical (http://www.caymanchem.com/pdfs/90080.pdf). Cannabidiol powder was dissolved in 95 % ethanol at a concentration of 10mg/ ml in our research pharmacy. This is similar to the stock solution prepared with Δ-9-THC by our research pharmacists. The stock solution prepared as above was filtered using a 0.2 um filter and a sample of this stock solution was submitted for assay, pH, pyrogenicity and sterility testing.

**Storage and Packaging:** The Cannabidiol solution was stored in closed bottles, protected from light at -20 degrees in the VA research pharmacy. Pharmacists delivered a single dose of this solution to study staff for administration to subjects.

**Stability and Quantitative analysis:** A sample of the stock solution was submitted for stability testing at 1week, 1 month, 3 months and 6 months by HPLC. the IND was amended accordingly to conduct stability testing at regular intervals.

**Table 1. Schedule of test days**

| Table 1: Schedule of Procedures | |
|---|---|
| **Time (minutes)** | **Procedure** |
| −60 | Confirmation of abstinence from caffeine, alcohol, drugs, medications<br>Urine drug screen, urine pregnancy test<br>Vital signs and placement of an intravenous line and pulse oximeter |
| −30 | **Behavioral assessments:**<br>PANSS<br>CADSS<br>VAS of feeling states<br>PSI<br>**Blood sampling**<br>**Vital signs** |
| 0 | IV placebo or active CBD followed by IV placebo or active THC over 20 minutes<br>Vital signs every 5 minutes until +30 |
| +15 | **Behavioral assessments**<br>PANSS<br>CADSS<br>VAS of feeling states<br>**Blood sampling** |
| +25<br><br>+50 | **Electroencephalography**<br>Resting State<br>P50 Auditory Sensory Gating<br>P300 Auditory Evoked Potential<br><br>**Cognitive Testing:**<br>Integ Neuro<br>CANTAB (MS , RVIP, SWM, SOC, DMTS) |
| +80 | **Behavioral assessments**<br>PANSS<br>CADSS<br>VAS of feeling states<br>**Blood sampling**<br>**Vital signs** |
| +120<br><br>+150 | **Vital Signs**<br><br>**Retrospective Behavioral assessments**<br>PANSS<br>CADSS<br>VAS of feeling states |

| Table 1: Schedule of Procedures | |
|---|---|
| **Time (minutes)** | **Procedure** |
| End of each day (+240) | **Behavioral assessments**<br>PANSS<br>CADSS<br>VAS of feeling states<br>PSI<br>Drug Liking Questionnaire<br>Mini Mental Status Examination<br>Field sobriety test<br>Physician evaluation<br>**Blood sampling**<br>**Vital Signs** |
| Next day, 1 and 3 months | Assessment for emergence of new psychiatric or medical problems<br>Assessment of drug use, desire, craving |

**Abbreviations:** CADSS: Clinician Administered Dissociative Symptoms Scale; HRS: Hallucinogen Rating Scale; PANSS: Positive and Negative Syndrome Scale; PSI: Psychotomimetic State s Inventory; VAS: Visual Analog Scale.
Blood sampling: for serum levels of THC and CBD, hormones and cannabinoid relevant molecules (CRM).

**Supplementary table 2:** Group differences between 18 subjects who participated only in phase-1 and 10 subjects who participated in both phases of the study

| Variable | Only Phase-1<br>n = 18 | Phase 1 and 2<br>n = 10 | Test statistic – p value |
|---|---|---|---|
| Age – median (IQR) | 26 (7.5) | 25 (10) | U = 65.5, p = 0.93 |
| Gender – female - n (%) | 8 (44.4%) | 4 (40%) | OR = 1.32 (0.21 – 8.91), p = 0.56 |
| Race – Caucasian: black: other (n) | 11:5:2 | 6:3:1 | P = 0.84 |
| Education – median (IQR) | 16 (2.5) | 15 (2) | U = 81.5, p = 0.25 |
| Cannabis – frequent user – n (%) | 4 (22.2%) | 0 (0) | OR = inf (0.36 – inf)<br>p = 0.26 |
| Smoker – n (%) | 3 (16.67%) | 2 (20%) | OR = 1.22 (0.08 – 13.65), p = >0.99 |
| Weight – pounds, median (IQR) | 162 (65.5) | 174 (32.2) | U = 67, p = 0.67 |
| BMI – median (IQR) | 25.4 (6.42) | 25.5 (5.9) | U = 63, p = 0.87 |
| NART – IQ – mean (SD) | 114 (7) | 110 (4.5) | U = 77.5, p = 0.27 |
| Psychosis proneness - median (IQR) | 17.5 (19) | 20.5 (10.25) | U = 48, p > 0.99 |
| SPQ score - median (IQR) | 4 (4) | 2.5 (6.75) | U = 58.5, p = 0.66 |

Supplementary figure 1 – study design

| Phase | Phase 1 | | | | Phase 2 | |
|---|---|---|---|---|---|---|
| **Days** | **Day 1** | **Day 2** | **Day 3** | **Day 4** | **Day 5** | **Day 6** |
| **Subjects** | **28** | | | | **10/28** | |
| **Treatment - randomization** | **Placebo / THC / CBD / CBD:THC (2:1)** | **Placebo / THC / CBD / CBD:THC (2:1)** | **Placebo / THC / CBD / CBD:THC (2:1)** | **Placebo / THC / CBD / CBD:THC (2:1)** | **CBD:THC (1:1) / CBD:THC (3:1)** | **CBD:THC (1:1) / CBD:THC (3:1)** |

Supplementary fig. 2A

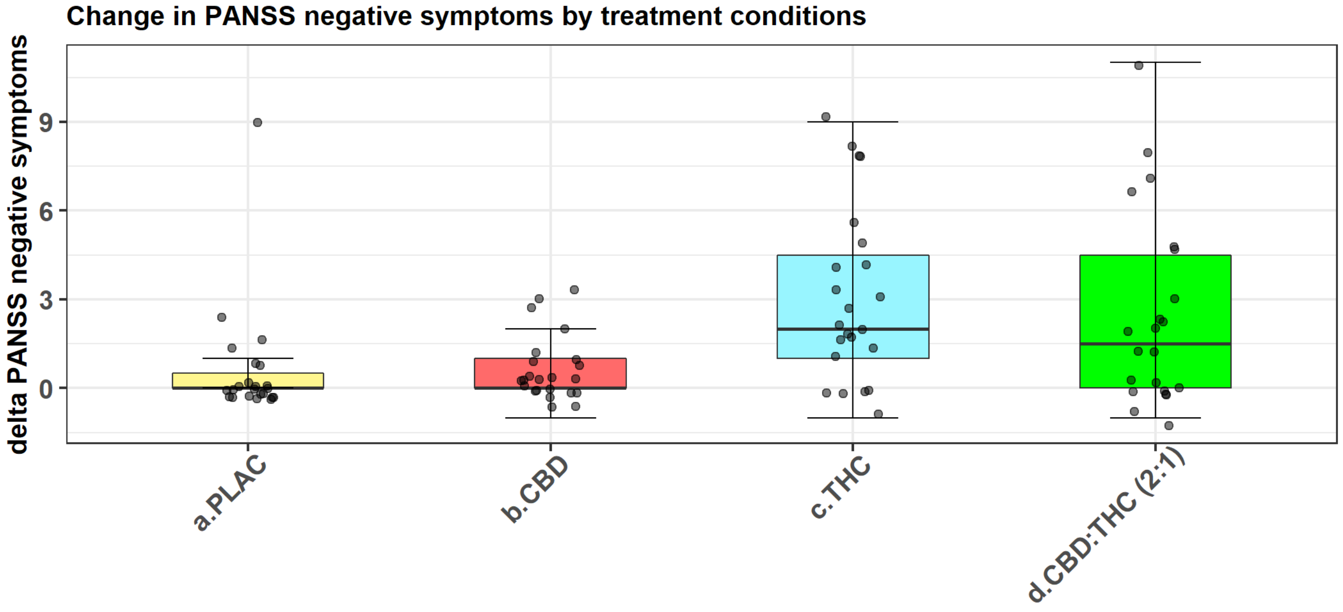


Supplementary fig. 2B

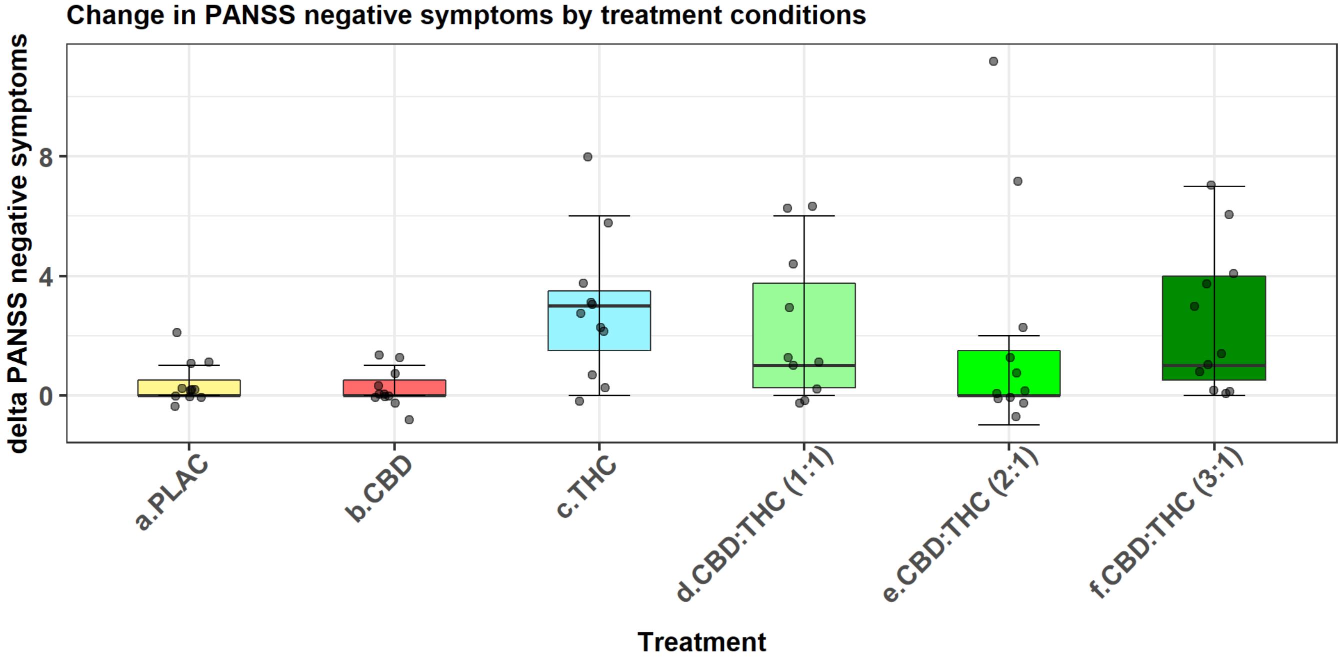

Supplementary fig. 3A

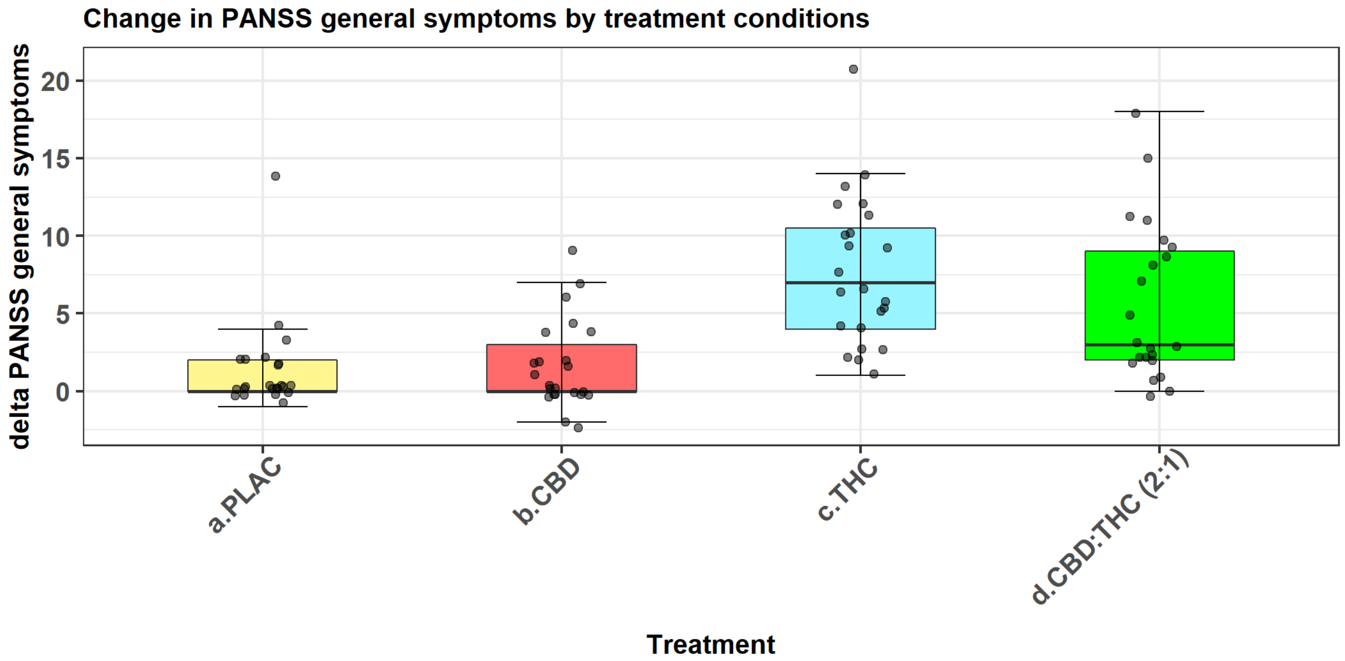


Supplementary fig. 3B

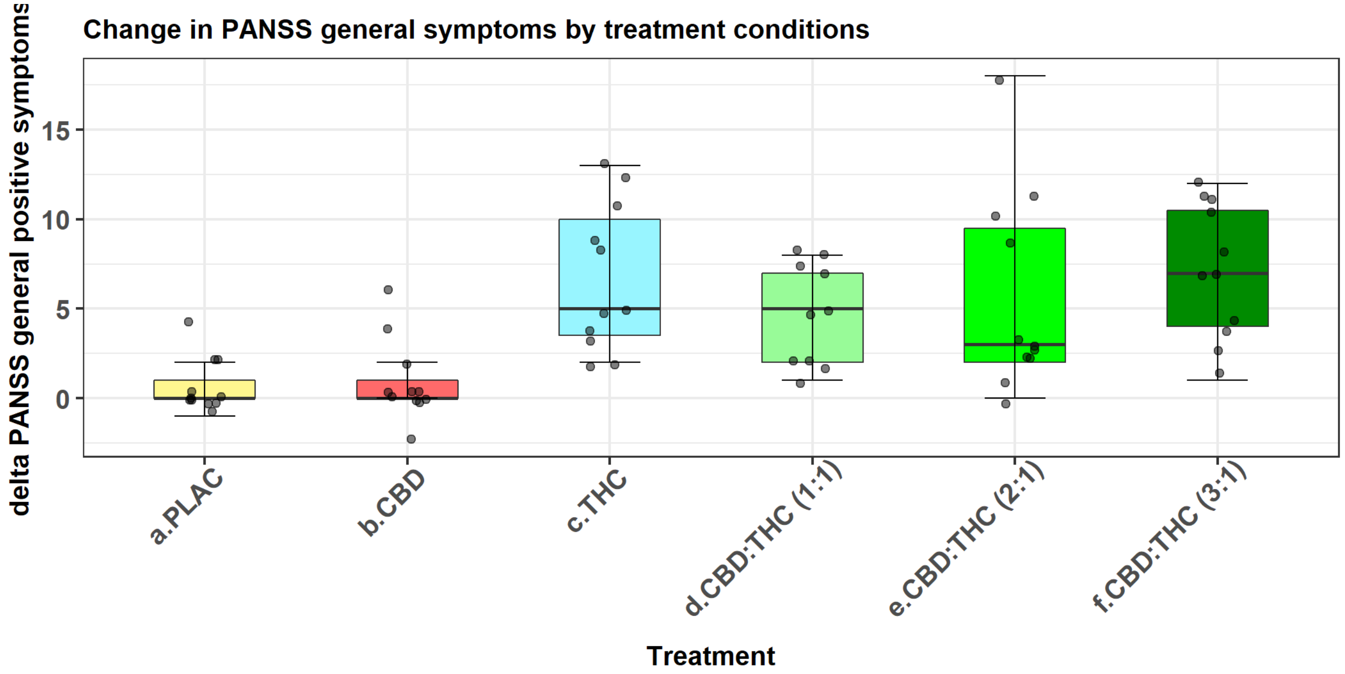

Supplementary fig. 4A

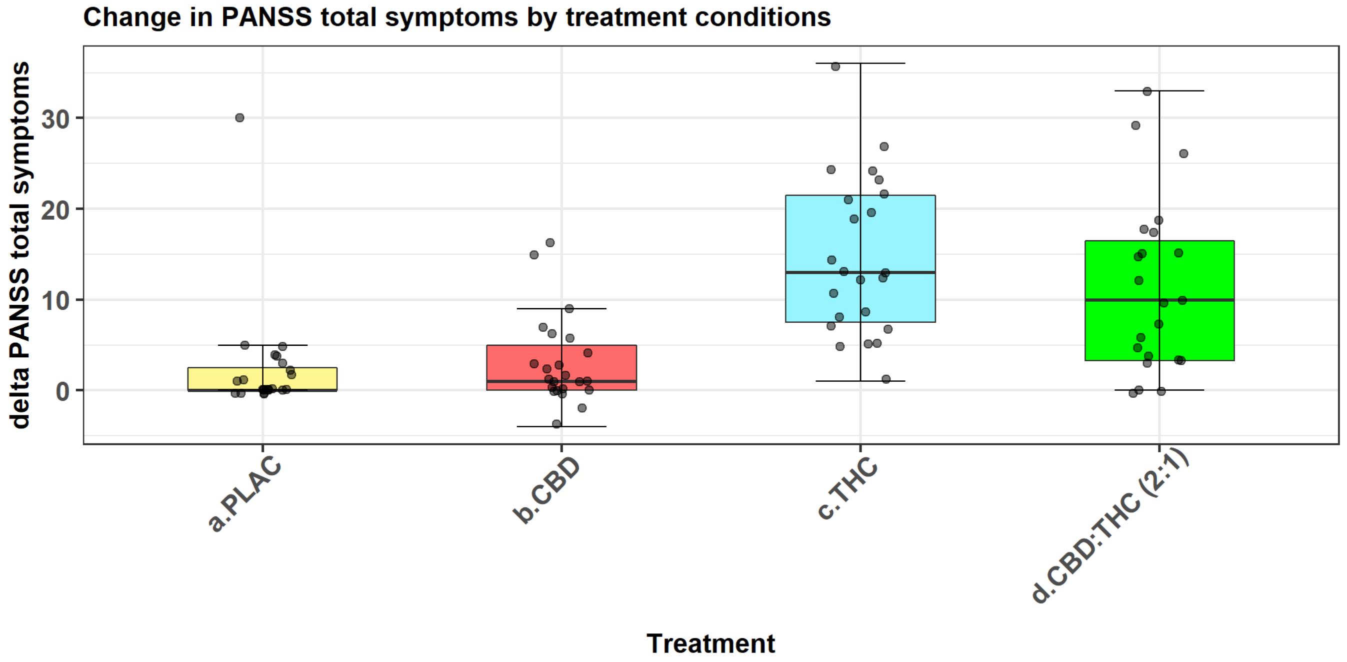


Supplementary fig. 4B

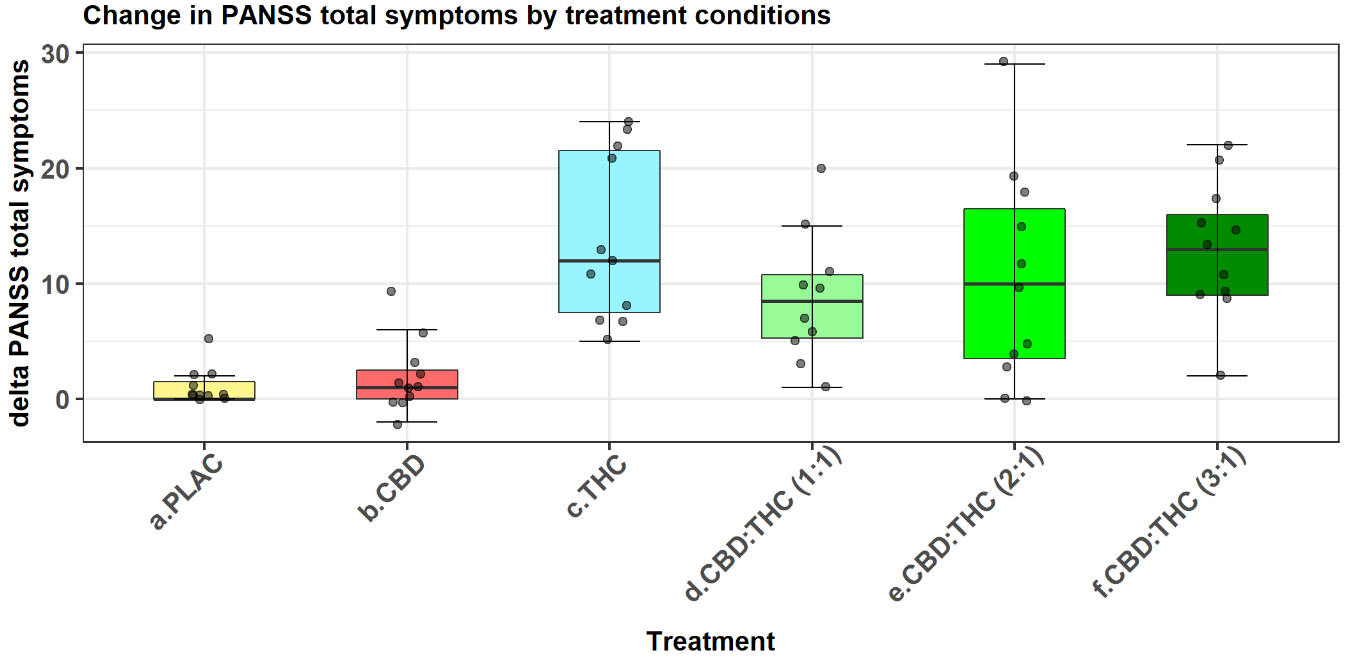

Supplementary fig. 5A

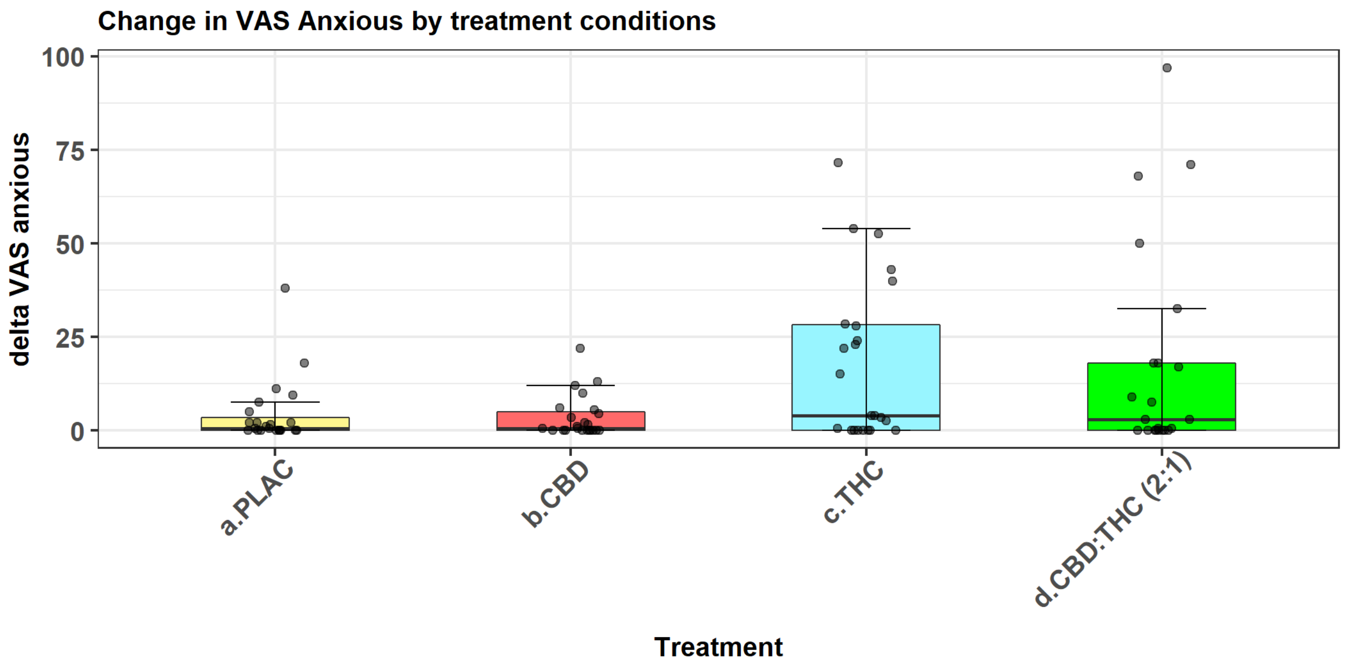


Supplementary fig. 5B

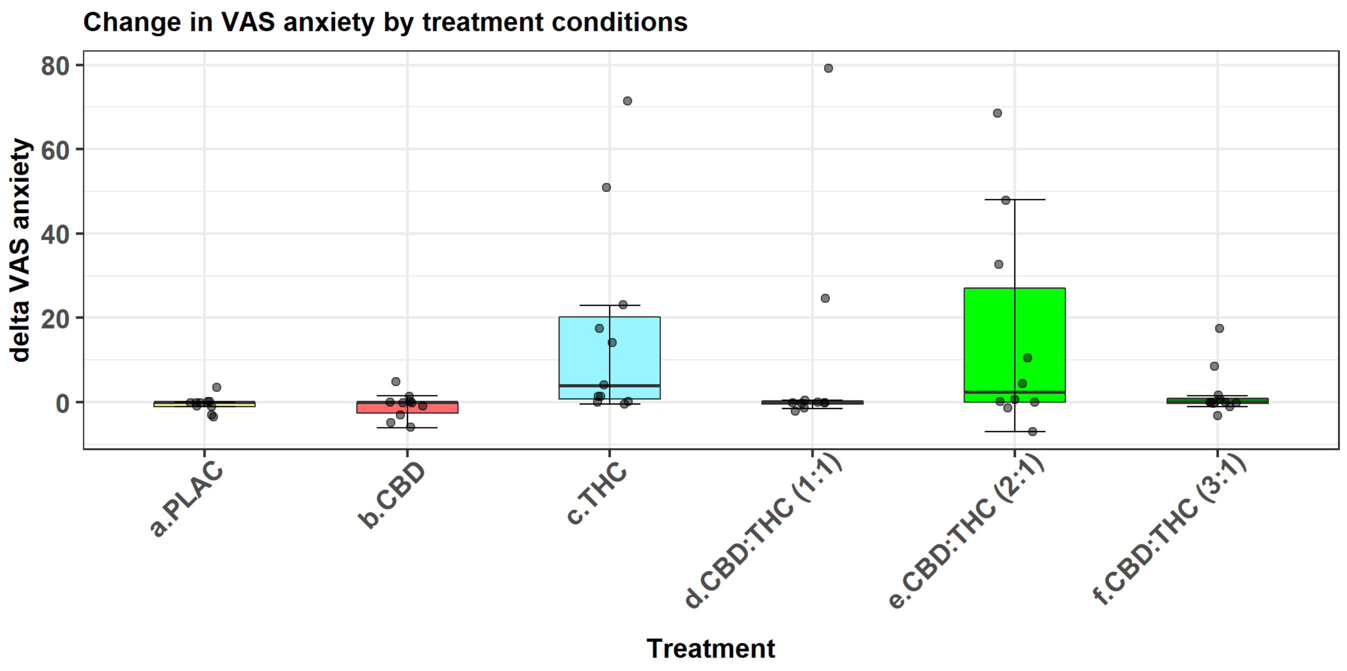

Supplementary fig. 6A

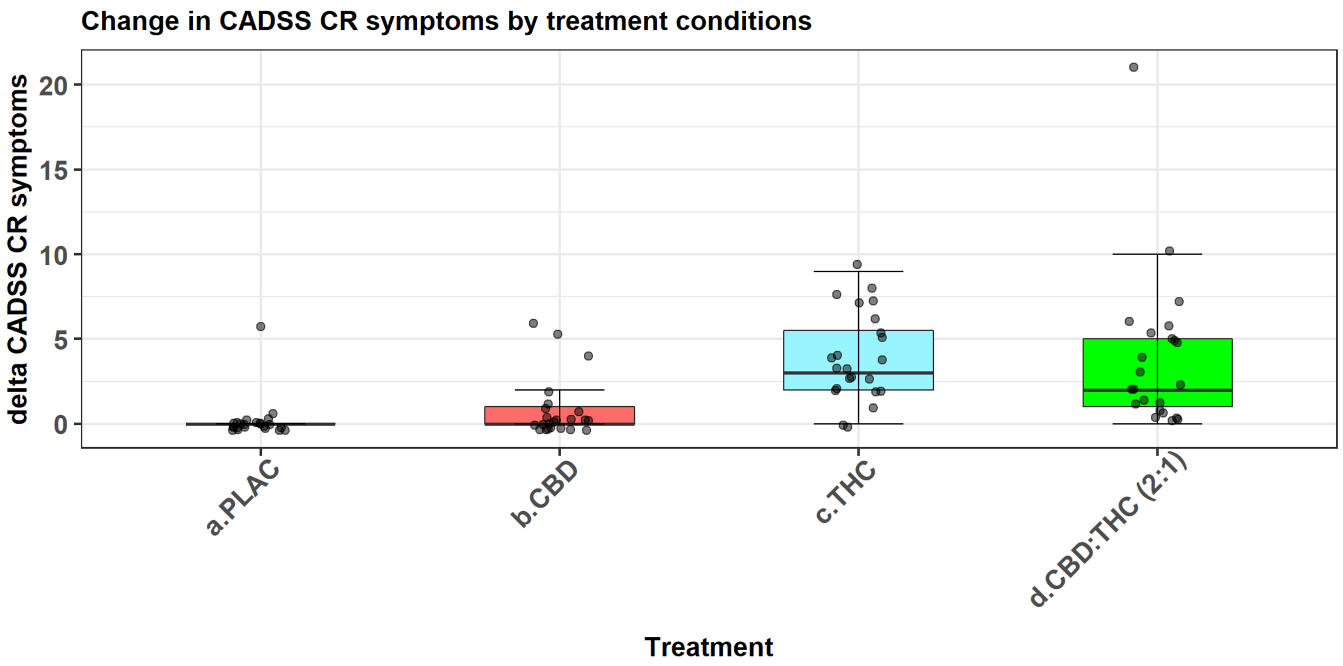


Supplementary fig. 6B

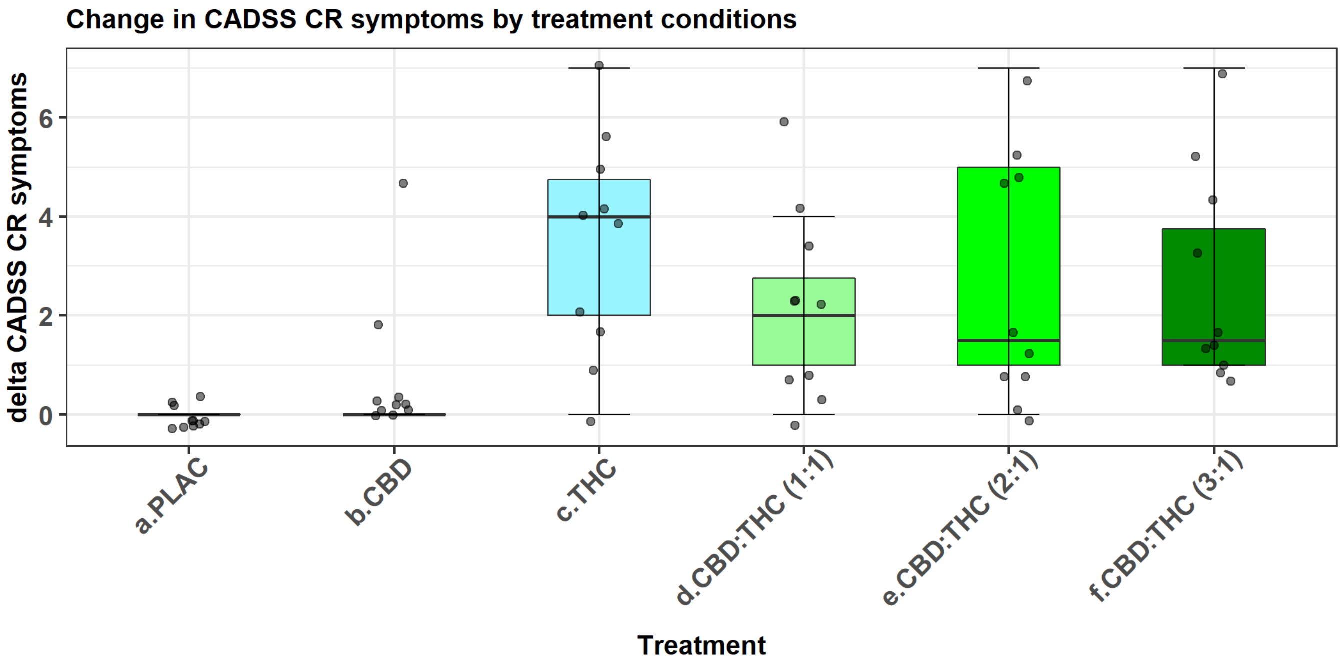

Supplementary fig. 7A

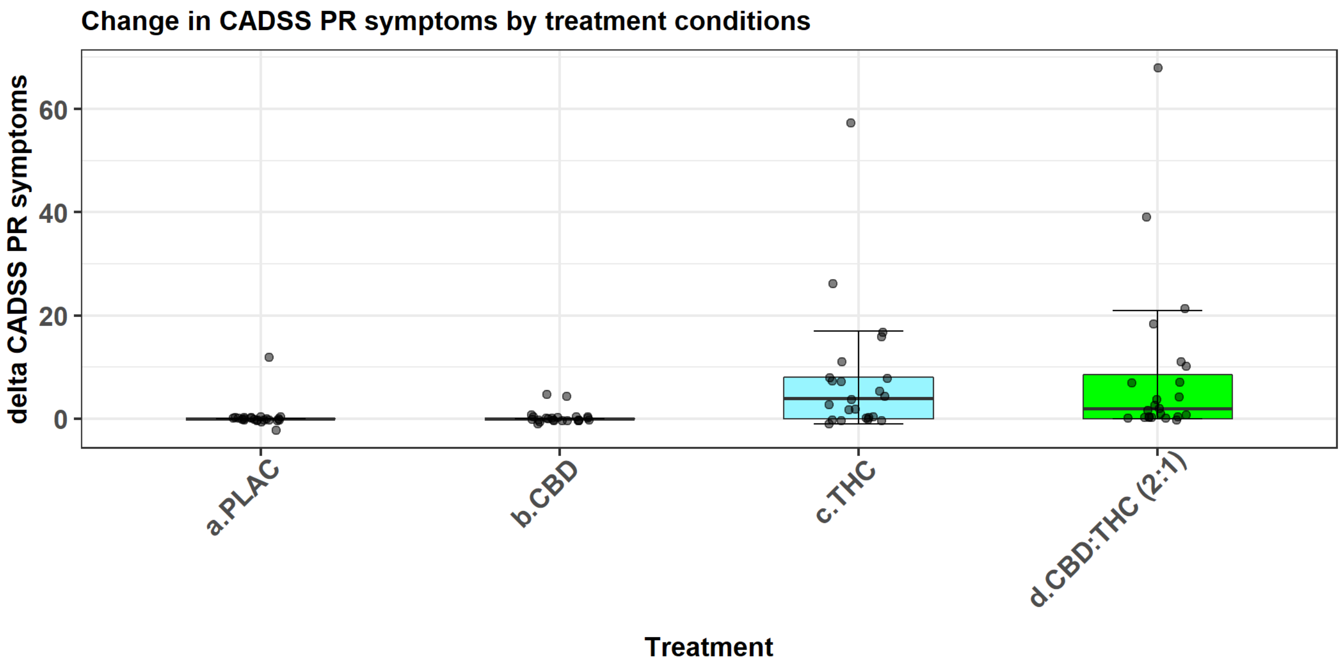


Supplementary fig. 7B

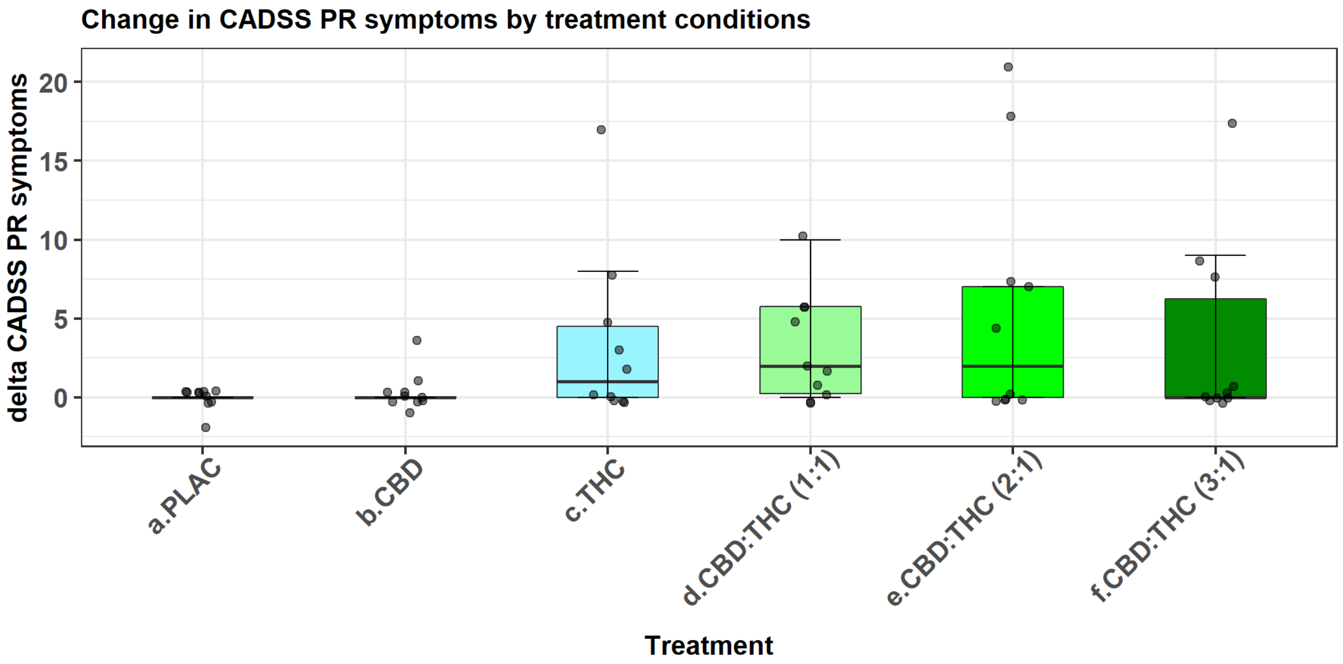

Supplementary fig. 8A

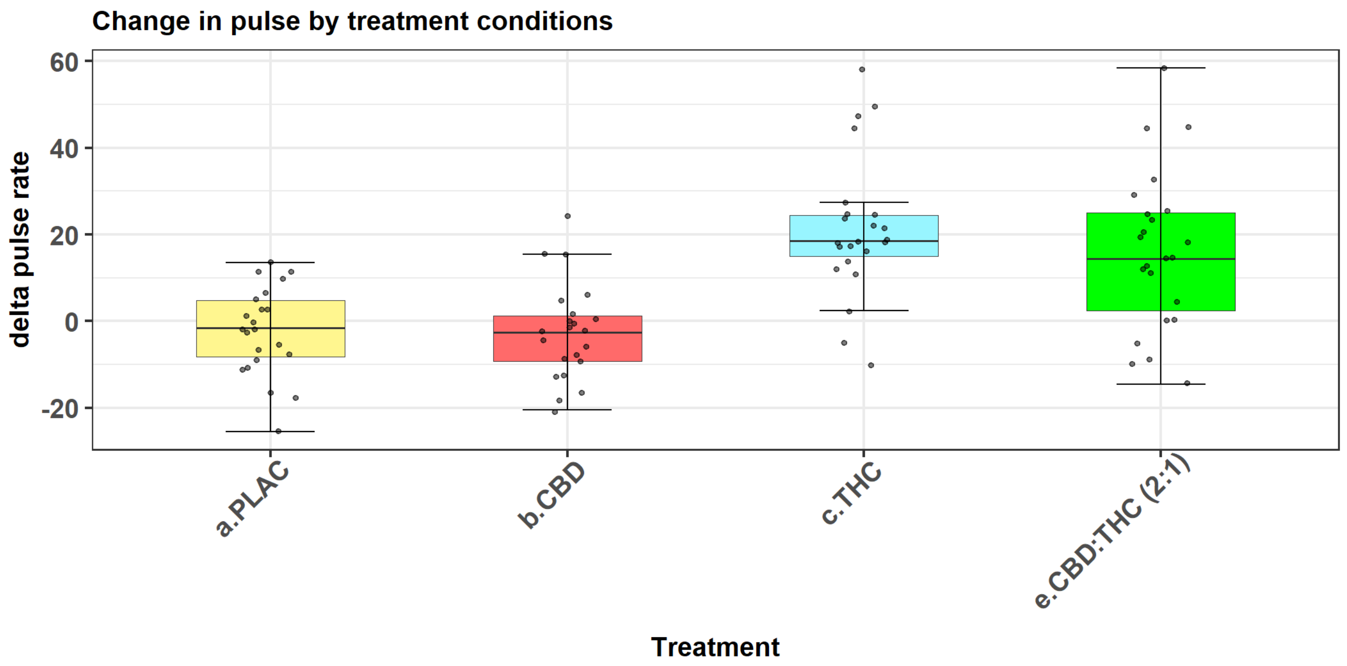


Supplementary fig. 8B

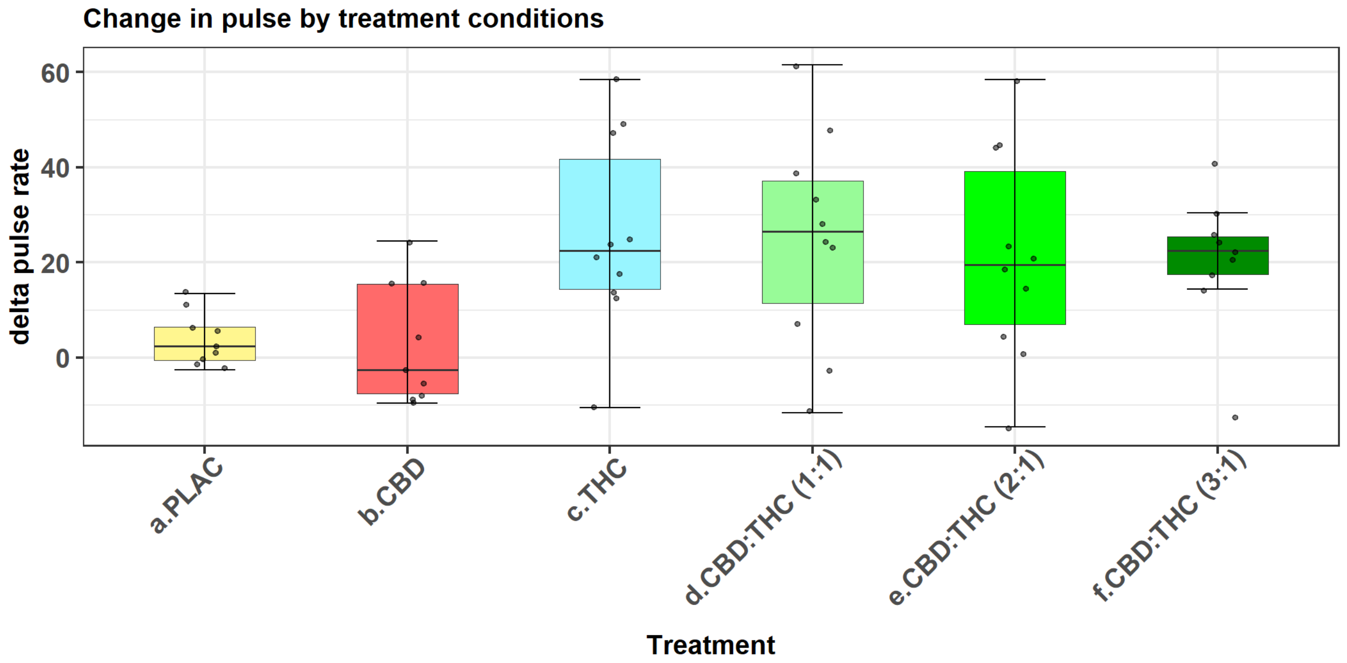
